# Procedure composition, age and race/color in gender-affirming surgical authorizations of the Brazilian public health system, 2014-2025

**DOI:** 10.64898/2026.11.23.26368723

**Authors:** Juliano dos Santos, Cristiane Maria Amorim Costa

## Abstract

Hospital admission authorizations from publicly financed health systems are increasingly used to describe gender-affirming surgery, although they record billed procedures whose feminizing or masculinizing direction can be inferred from the code, without identifying people, self-reported gender identity or clinical need. This nationwide descriptive study examined the authorizations for gender-affirming surgery processed by the Brazilian Unified Health System from 2014 to 2025, which were selected from 3,888 public monthly state files of the Hospital Information System with a historically versioned classification of procedure codes. Of the 1,375 selected authorizations, 711 corresponded to directly feminizing procedures, 428 to directly masculinizing procedures and 236 to a complementary procedure that was analyzed separately. The feminizing share of the directly classified authorizations decreased from 81.7% (196/240) in 2014-2018 to 72.2% (213/295) in 2019-2023 and to 50.0% (302/604) in 2024-2025, proportions that describe composition within periods of unequal length and cannot be read as population rates. In 2024-2025, bilateral mastectomy accounted for 264 of the 302 masculinizing authorizations (87.4%), whereas bilateral breast reconstruction with silicone implants accounted for 118 of the 302 feminizing authorizations (39.1%). The median recorded age was 35 years in feminizing and 30 years in masculinizing authorizations over the whole period, and 34 and 29 years when the comparison was restricted to 2024-2025. The brown (parda) race/color category was recorded in 42.6% (303/711) of feminizing and 61.0% (261/428) of masculinizing authorizations, in a comparison pooled over the whole period and not adjusted for period or procedure. The number of hospitals habilitated for these surgeries remained at five from 2014 to 2022 and doubled in 2023. All aggregate results were reproduced by an independent pipeline that re-read every original public file. It was observed that the equal numbers of feminizing and masculinizing authorizations in the most recent period resulted from markedly different procedure portfolios, which suggests that the monitoring of publicly funded gender-affirming surgery may benefit from procedure-specific indicators and versioned code definitions, while questions of access and equity would require person-level and patient-reported data.

## Introduction

Gender-affirming care in the Brazilian Unified Health System (Sistema Único de Saúde, SUS) is organized through the Transsexualizing Process, a national policy that was redefined and expanded in 2013 with outpatient and hospital components, accredited specialized services and specific procedure codes, and that set the minimum age of 21 years and two years of prior multiprofessional follow-up for surgical procedures [1]. Brazilian studies have examined the implementation of this policy, its advances and challenges, the itineraries that users follow to reach specialized services and the trajectory of health policy in gender transition processes [2–5], and, since rules of eligibility, accreditation and financing changed while the policy was in force, hospital counts of gender-affirming surgery need to be read in the light of these rules.

Hospital and billing records have been used in other countries to estimate the volume of gender-affirming surgery and its distribution by type of procedure, age and region [6–10], with national samples in the United States in which breast and chest procedures were the most frequent group [6,7]. In Germany, hospital quality reports described the growth of masculinizing and feminizing genital surgery over almost two decades [9]. It should be noted, however, that the correspondence between administrative codes and self-reported gender identity or sex assigned at birth has required specific validation studies [11,12], whose estimates of accuracy cannot be transposed to a classification based on the procedure table of the SUS.

In Brazil, a conference abstract based on the Hospital Information System reported a marked increase in bilateral mastectomy in the gender-affirming process from 2023 onwards [13], and a nationwide conference analysis of the same system for January 2014 to November 2025 described hospitalization volume, procedure types, length of stay and financial indicators [14]. Administrative data of the SUS have also been used to describe long-term changes in the composition of publicly financed surgery in other fields [15], while studies with patient-reported data have documented experiences of access to transition-related care and the role of discrimination and race in the health of sexual and gender diverse people, aspects that billing records cannot capture [16,17]. In this context, it was observed that no previous analysis had jointly examined the direction of the coded procedures under historical eligibility rules, the age and race/color recorded in the authorizations and the procedure composition behind the aggregate counts.

The present study aimed to describe the gender-affirming surgical authorizations processed by the SUS from 2014 to 2025 according to procedure direction, period, recorded age and recorded race/color, and to examine whether similar recent numbers of feminizing and masculinizing authorizations corresponded to similar procedure compositions and demographic profiles. This aim was achieved at the level of administrative episodes, and the limits of this unit of analysis for inferences about people, demand and access are discussed below.

## Materials and methods

### Study design, data source and period

A nationwide, retrospective and descriptive study was conducted with routinely collected hospital data, reported in accordance with the RECORD extension of the STROBE statement (S3 Checklist) [18]. The data source was the reduced file (RD) of the Hospital Information System of the SUS (Sistema de Informações Hospitalares, SIH/SUS), which the Department of Informatics of the SUS (DATASUS) disseminates publicly as one compressed file per federative unit and processing month [19]. The study covered the processing months from January 2014 to December 2025, which corresponds to 27 federative units over 144 months, or 3,888 source files. The unit of analysis was the hospital admission authorization (Autorização de Internação Hospitalar, AIH), which records a billed hospitalization and its performed procedure, so that one person may contribute more than one authorization and the processing month may differ from the date of surgery.

### Selection and classification of procedures

Authorizations were selected when the performed-procedure field (PROC_REA) contained a gender-affirming surgical code that was valid in the processing month, according to a historical crosswalk of the procedure table of the SUS (SIGTAP) compiled from monthly versions of the table and from the corresponding ordinances (S1 Appendix). The directly feminizing group comprised thyroplasty (0404010563), the feminizing genital procedure (0409050148) and bilateral breast reconstruction with silicone implants (0410010200), and the directly masculinizing group comprised hysterectomy with bilateral adnexectomy and colpectomy (0409060291), bilateral mastectomy (0410010197), the masculinizing genital procedure (0409070319) and neophalloplasty (0409050164). The code for complementary procedures (0409050130), which the crosswalk classifies as predominantly feminizing but which has lower directional specificity, was analyzed as a separate group and added to the feminizing group in a sensitivity analysis, and the code for sequential procedures (0415020085) was not assigned to either direction and had no selected records. The three feminizing codes, hysterectomy, bilateral mastectomy and the complementary code were valid throughout the study period, whereas the masculinizing genital procedure entered the table in July 2019, the sequential procedures in January 2023 and neophalloplasty in January 2024, with its first monthly observation in the table in February 2024, and these windows were applied in the selection.

The labels feminizing and masculinizing describe the direction of the coded procedure and were not used as indicators of self-identified gender, sex assigned at birth, clinical indication or a validated identification of transgender people, an approach consistent with the SAGER recommendations on the reporting of sex and gender [20–22]. A principal diagnosis in category F64 of the International Classification of Diseases was not an inclusion criterion and was examined only as a descriptive flag. No validated person-level identifier was available, and authorizations were therefore not deduplicated into individuals. The habilitations accepted for each procedure were taken from the monthly SIGTAP tables [23]. The establishments habilitated to perform these surgeries were counted in the public habilitation files of the CNES for December of each year, considering codes 30.01, the specialized unit of the Transsexualizing Process created in 2008, and 30.03, pre- and postoperative care and surgery, the two codes accepted for these procedures throughout the period, with the outpatient code 30.02 counted separately [24].

### Variables

The share of feminizing authorizations among the directly classified authorizations was calculated as F/(F+M) for the processing periods 2014-2018, 2019-2023 and 2024-2025, which cover five, five and two years, and multi-year periods were adopted because the masculinizing group had only 44 authorizations in 2014-2018, which implies annual counts below 10 in at least one of those years. Age was taken from the age field when the age unit indicated years. Race/color was taken from the SIH/SUS field [19], coded as white (branca), black (preta), brown (parda), Asian (amarela) and Indigenous (indígena), plus code 99 for not informed, and it was not possible to establish whether each entry had been self-declared. For the period 2024-2025, the two breast and chest procedures, bilateral mastectomy and bilateral breast reconstruction with silicone implants, were described by their counts, by their share of the respective directed group and by recorded age and race/color, considering that these operations have different indications, eligibility requirements and care pathways and cannot be treated as equivalent interventions. Counts by procedure were also reported for the three periods, with hysterectomy, the masculinizing genital procedure and neophalloplasty pooled to avoid cells below 10.

### Statistical analysis

All analyses were descriptive, and the study population comprised all selected authorizations, so that no sampling or sample size calculation was involved. Counts and percentages were reported with numerators and denominators, and age was summarized by median, first quartile (Q1) and third quartile (Q3), calculated by linear interpolation, which is the default method of pandas and NumPy. Race/color percentages used all authorizations of each directed group as the denominator, code 99 remained in the denominator and no values were imputed. Because authorizations from the same person and from the same establishment may be correlated, hypothesis tests and confidence intervals that assume independent observations were not calculated, and no correction for multiple comparisons was therefore required. To reduce the influence of pooling different calendar periods, the age comparison was repeated for 2024-2025, a period in which both groups contained the same number of authorizations. Records with implausible recorded ages were retained in the main series, and a sensitivity analysis restricted to authorizations with recorded age of 18 years or more was carried out. Race/color was compared between the whole directed groups only in the pooled 2014-2025 series and was described for 2024-2025 only within the two breast and chest procedures, and analyses adjusted for period, procedure or establishment were not performed. The independent replay was written in Python 3.13 with the datasus-dbc 0.1.3 and dbfread 2.0.7 packages, the figures were drawn in R 4.5.2 with ggplot2 4.0.3, and the code, the crosswalk and the aggregate results are available in Zenodo (https://doi.org/10.5281/zenodo.24117844).

### Data quality and reproducibility

The analytic extract comprised 1,375 authorizations and 59 variables and was checked against archived processing logs, which recorded 119,005,606 source rows for 2014-2023, 693 selected authorizations for 2014-2023 and 682 for 2024-2025, with no exact duplicate rows. The first processing month with a selected record was June 2014, and a specific check of January to May 2014 found no target procedures in the source files. All aggregate results reported in this article were subsequently reproduced by an independent pipeline that downloaded and decoded the 3,888 original files, applied the historical crosswalk without using the analytic extract as input and passed all 14 prespecified comparisons, which covered the totals by group and period, the age quartiles, the race/color categories and the breast and chest subgroups. The municipality of the establishment recorded in the SIH/SUS (MUNIC_MOV) agreed with the municipality of the same establishment in the historical register of the National Registry of Health Establishments (CNES) for all 1,139 directly classified authorizations, matched by establishment and processing month, and the managing municipality field (UF_ZI) was not used as a proxy for the location of care [19]. Data provenance, the crosswalk, the selection flow and the checks are detailed in S1 Appendix and in the analysis protocol documented retrospectively in S2 Protocol.

### Ethics statement

The study used only SIH/SUS, SIGTAP and CNES files that DATASUS disseminates publicly without individual authorization, with no recruitment of or contact with participants, no intervention and no access to restricted or identified records. For the results reported here, the SIH/SUS files were accessed for research purposes on 9 and 10 November 2026, when the independent reproduction downloaded all 3,888 files, and the SIGTAP tables and CNES habilitation files on 16 November 2026, while the analytic extract had been built earlier from the same public files. The public files contain individual records without names, with indirect identifiers such as date of birth, sex and municipality of residence, and the authors had no access to names or other direct identifiers and did not use these fields to identify persons. Article 26, item II, of Resolution No. 674/2022 of the Brazilian National Health Council exempts from review by the research ethics committee system (CEP/Conep) research that uses information of public access under Law No. 12,527/2011 [25], and the study was therefore not submitted to an ethics committee and did not require informed consent. This exemption derives from the regulation itself, and no waiver was requested from a committee. No attempt was made to identify individuals or to link authorizations to persons, the event-level extract derived by the authors is not redistributed and cells with fewer than 10 authorizations in combinations of procedure, period, place and social attributes were not reported, a threshold adopted as an internal screening rule that does not by itself guarantee anonymization.

### Use of artificial intelligence tools

Anthropic Claude (Claude Opus 5.5) was used in the final revision to verify each reference against Crossref records and official sources. The tool was not used as a data source or to produce results. All numerical results were checked against the outputs of the independent replay and of the analytic extract, and the authors reviewed the entire text and are responsible for the study design, analytic decisions, interpretation and final content.

## Results

### Selected authorizations and composition by period

Of the 1,375 selected authorizations, 711 (51.7%) were directly feminizing, 428 (31.1%) directly masculinizing and 236 (17.2%) complementary, with 158 complementary authorizations in 2014-2023 and 78 in 2024-2025, and 1,343 of the 1,375 authorizations (97.7%) had a principal diagnosis in category F64. The annual number of selected authorizations increased from 24 in 2014 to 112 in 2019, decreased to 39 in 2020 and 38 in 2021 and reached 126 in 2023, 272 in 2024 and 410 in 2025 (Fig 1). The establishments habilitated for these surgeries remained at five, in five federative units, from 2014 to 2022, increased to 10, in seven units, in 2023 and 2024 and reached 12, in eight units, in 2025, whereas outpatient habilitations increased from one to 34 (S1 Appendix). From the September 2023 competency, the SIGTAP also accepted for these procedures the habilitation of the National Program for Reducing Elective Surgery Queues and, in December 2025, three habilitations of the Agora Tem Especialistas program. Among the 1,139 directly classified authorizations, the feminizing share was 81.7% (196/240) in 2014-2018, 72.2% (213/295) in 2019-2023 and 50.0% (302/604) in 2024-2025, when each group contributed 302 authorizations (Table 1). When the complementary authorizations were added to the feminizing group, the feminizing share was 81.8% (567/693) in 2014-2023 and 55.7% (380/682) in 2024-2025.

**Fig 1.**
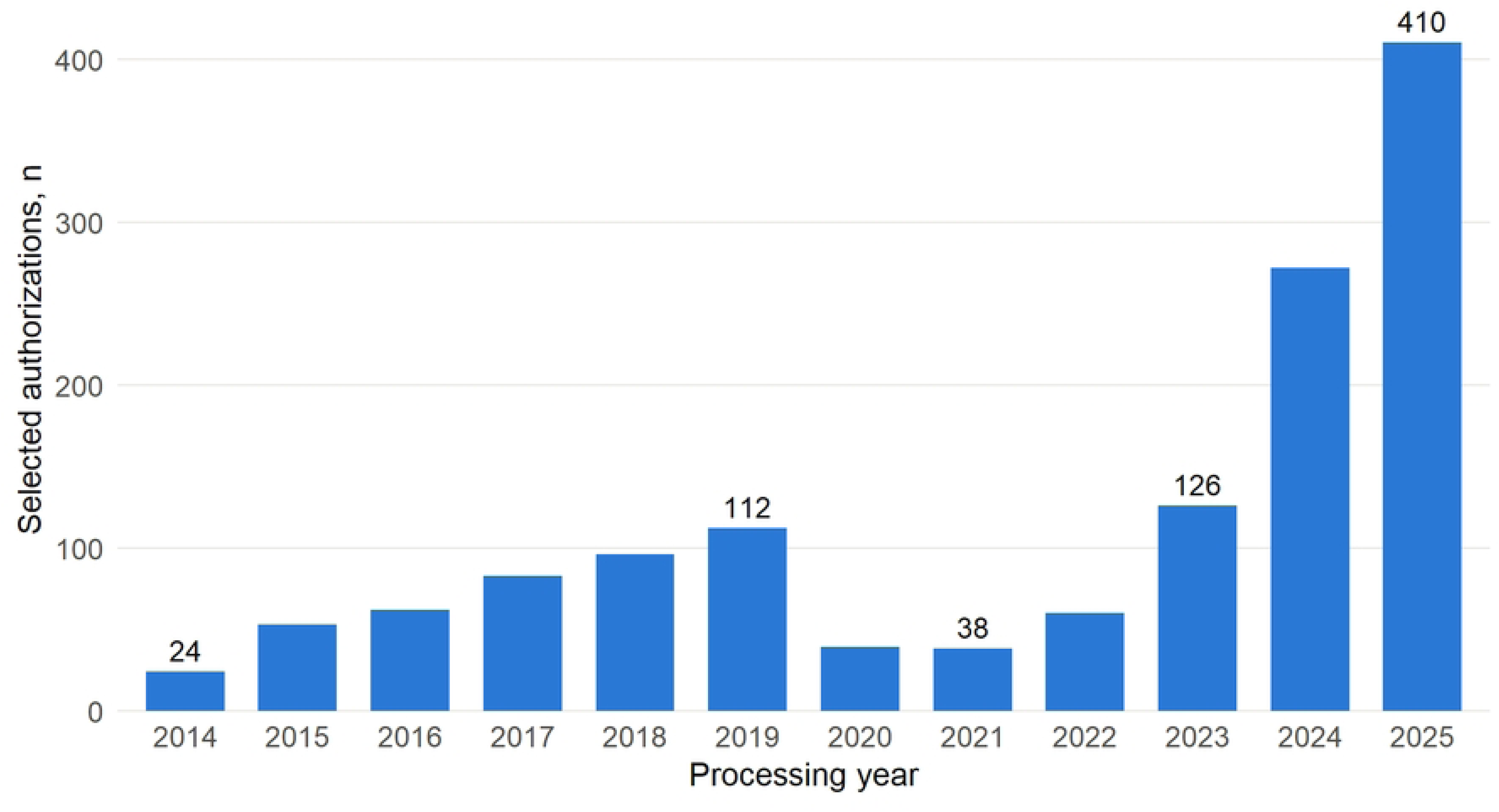
Selected gender-affirming surgical authorizations by processing year, Brazil, 2014-2025. The unit is the hospital admission authorization (AIH), and the totals include the directly feminizing, directly masculinizing and complementary groups.

**Table 1.**
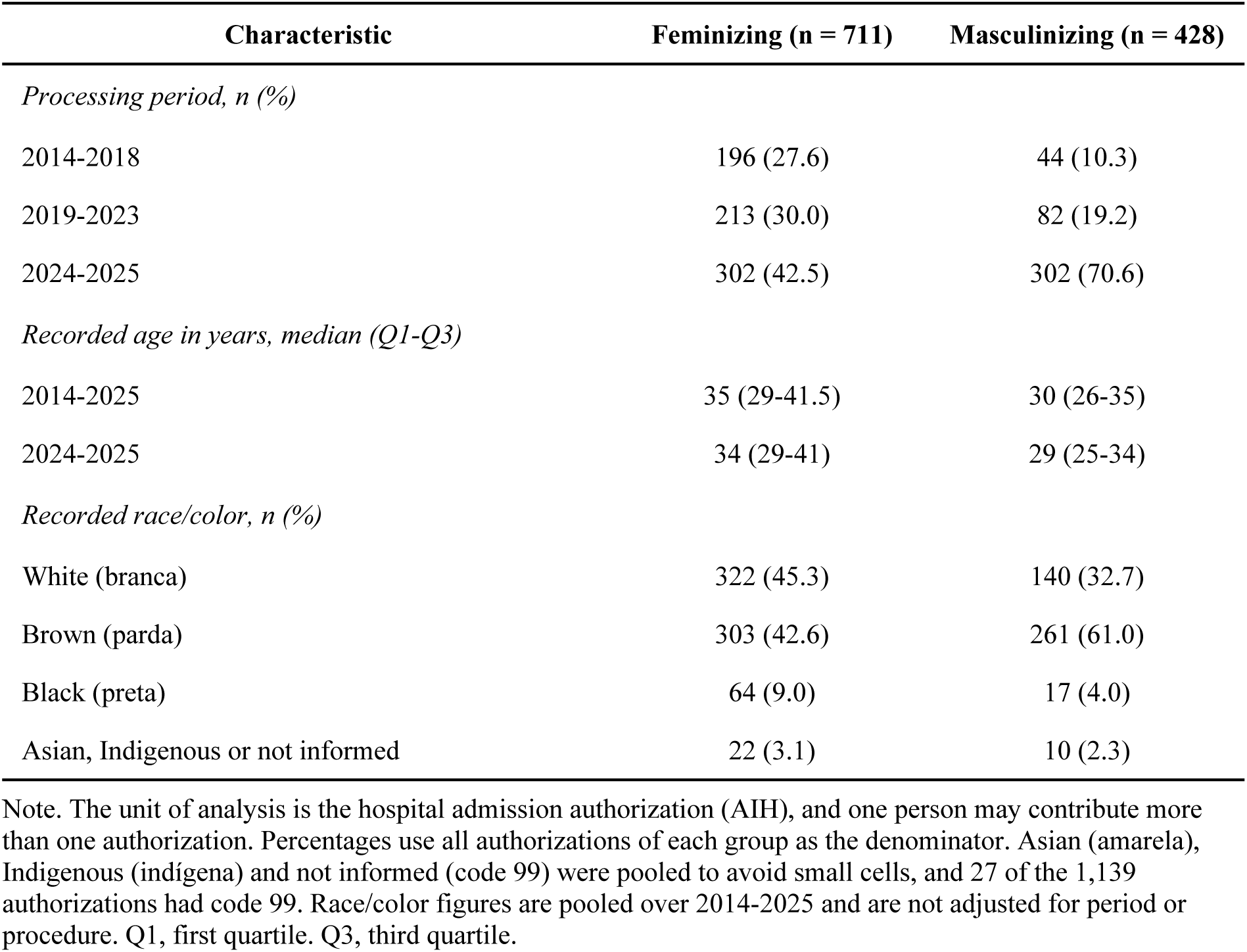
Characteristics of the directly classified authorizations by procedure direction, Brazil, 2014-2025.

| **Characteristic** | **Feminizing (n = 711)** | **Masculinizing (n = 428)** |
|---|---|---|
| *Processing period, n (%)* |  |  |
| 2014-2018 | 196 (27.6) | 44 (10.3) |
| 2019-2023 | 213 (30.0) | 82 (19.2) |
| 2024-2025 | 302 (42.5) | 302 (70.6) |
| *Recorded age in years, median (Q1-Q3)* |  |  |
| 2014-2025 | 35 (29-41.5) | 30 (26-35) |
| 2024-2025 | 34 (29-41) | 29 (25-34) |
| *Recorded race/color, n (%)* |  |  |
| White (branca) | 322 (45.3) | 140 (32.7) |
| Brown (parda) | 303 (42.6) | 261 (61.0) |
| Black (preta) | 64 (9.0) | 17 (4.0) |
| Asian, Indigenous or not informed | 22 (3.1) | 10 (2.3) |

Note. The unit of analysis is the hospital admission authorization (AIH), and one person may contribute more than one authorization. Percentages use all authorizations of each group as the denominator. Asian (amarela), Indigenous (indígena) and not informed (code 99) were pooled to avoid small cells, and 27 of the 1,139 authorizations had code 99. Race/color figures are pooled over 2014-2025 and are not adjusted for period or procedure. Q1, first quartile. Q3, third quartile.

### Composition by procedure

By procedure, the feminizing genital procedure accounted for 151, 138 and 150 authorizations in 2014-2018, 2019-2023 and 2024-2025, whereas breast reconstruction increased from 33 and 44 to 118 and bilateral mastectomy from 23 and 46 to 264 (Table 2 and Fig 2). Thyroplasty accounted for 12, 31 and 34 authorizations, and hysterectomy, the masculinizing genital procedure and neophalloplasty, reported together, for 21, 36 and 38. Breast and chest surgery therefore represented 16.8% (33/196) of feminizing authorizations in 2014-2018 and 39.1% (118/302) in 2024-2025, and 52.3% (23/44) and 87.4% (264/302) of masculinizing authorizations in the same periods.

**Table 2.** Selected authorizations by procedure and processing period, Brazil, 2014-2025.

| **Procedure (SIGTAP code)** | **2014-2018** | **2019-2023** | **2024-2025** | **Total** |
|---|---|---|---|---|
| *Directly feminizing* |  |  |  |  |
| Thyroplasty (0404010563) | 12 | 31 | 34 | 77 |
| Feminizing genital procedure (0409050148) | 151 | 138 | 150 | 439 |
| Bilateral breast reconstruction with silicone implants (0410010200) | 33 | 44 | 118 | 195 |
| *Directly masculinizing* |  |  |  |  |
| Bilateral mastectomy (0410010197) | 23 | 46 | 264 | 333 |
| Hysterectomy, masculinizing genital procedure or neophalloplasty (0409060291, 0409070319, 0409050164) | 21 | 36 | 38 | 95 |
| *Complementary* |  |  |  |  |
| Complementary procedures (0409050130) | 78 | 80 | 78 | 236 |
| Total | 318 | 375 | 682 | 1,375 |

Note. Counts of selected hospital admission authorizations (AIH) from the national summaries of the independent reproduction. Hysterectomy, the masculinizing genital procedure (valid from July 2019) and neophalloplasty (valid from January 2024) were pooled to avoid cells below 10. The periods cover five, five and two years, and their totals are not comparable as annual rates.

**Fig 2.**
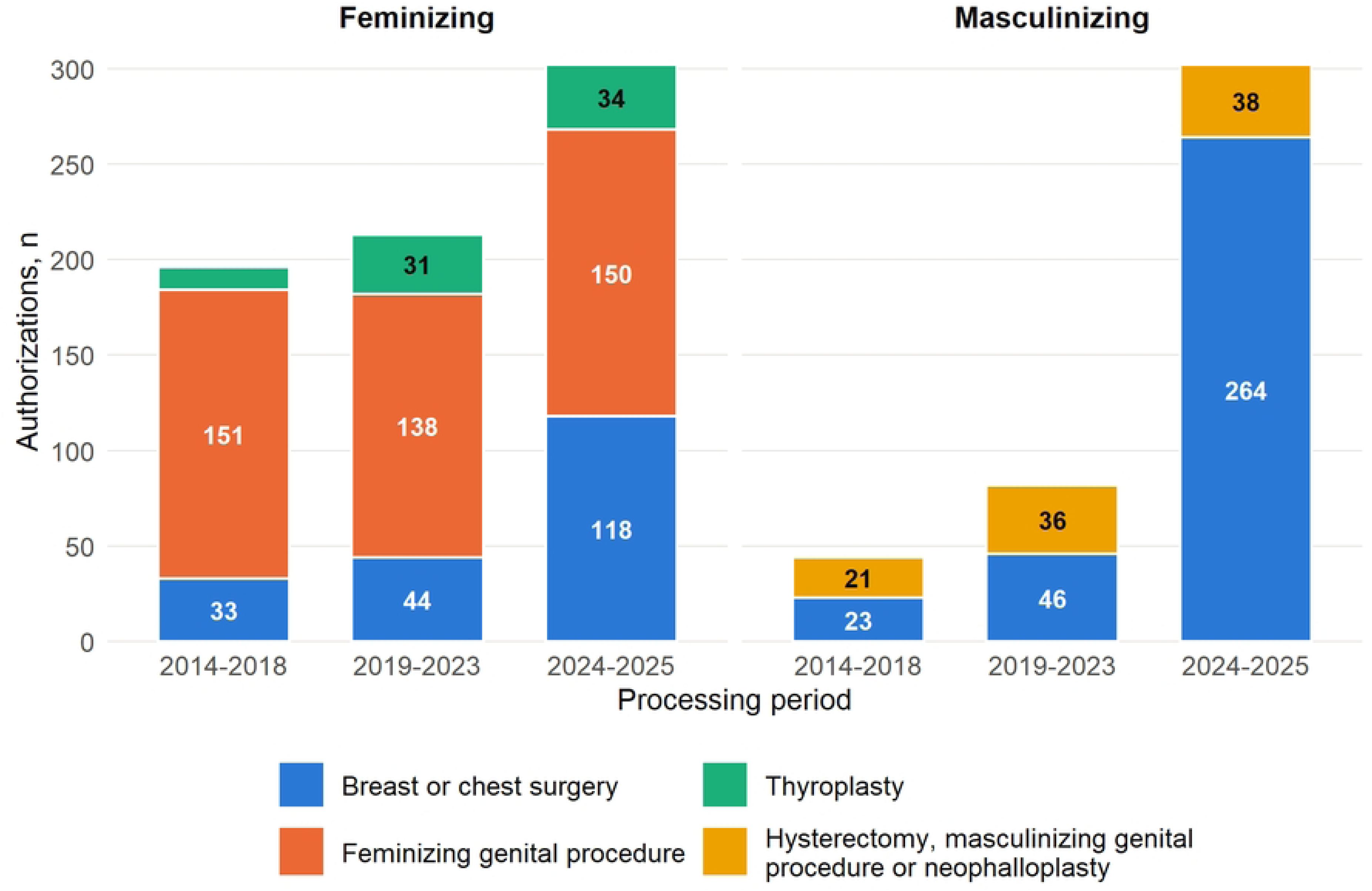
Selected authorizations by procedure and processing period in the directly feminizing and directly masculinizing groups, Brazil, 2014-2025. Breast or chest surgery corresponds to bilateral breast reconstruction with silicone implants in the feminizing group and to bilateral mastectomy in the masculinizing group. Segments below 20 authorizations are not labeled, and their values are given in Table 2.

### Recorded age and race/color

The median recorded age was 35 years (Q1-Q3, 29-41.5) in feminizing and 30 years (26-35) in masculinizing authorizations over 2014-2025, and 34 years (29-41) and 29 years (25-34) in 2024-2025 (Table 1 and Fig 3). Fewer than 10 authorizations had recorded ages below 21 years, all of them processed before 2024 and including values incompatible with any surgical indication, and the restriction to recorded ages of 18 years or more did not change the medians of either group. An informative race/color code was present in 1,112 of the 1,139 directly classified authorizations (97.6%), and all 27 authorizations with code 99 were processed before April 2023. The brown category was recorded in 303 of the 711 feminizing (42.6%) and in 261 of the 428 masculinizing authorizations (61.0%), the black category in 64 (9.0%) and 17 (4.0%), and the white category in 322 (45.3%) and 140 (32.7%), respectively, so that black and brown together accounted for 367 (51.6%) feminizing and 278 (65.0%) masculinizing authorizations.

**Fig 3.**
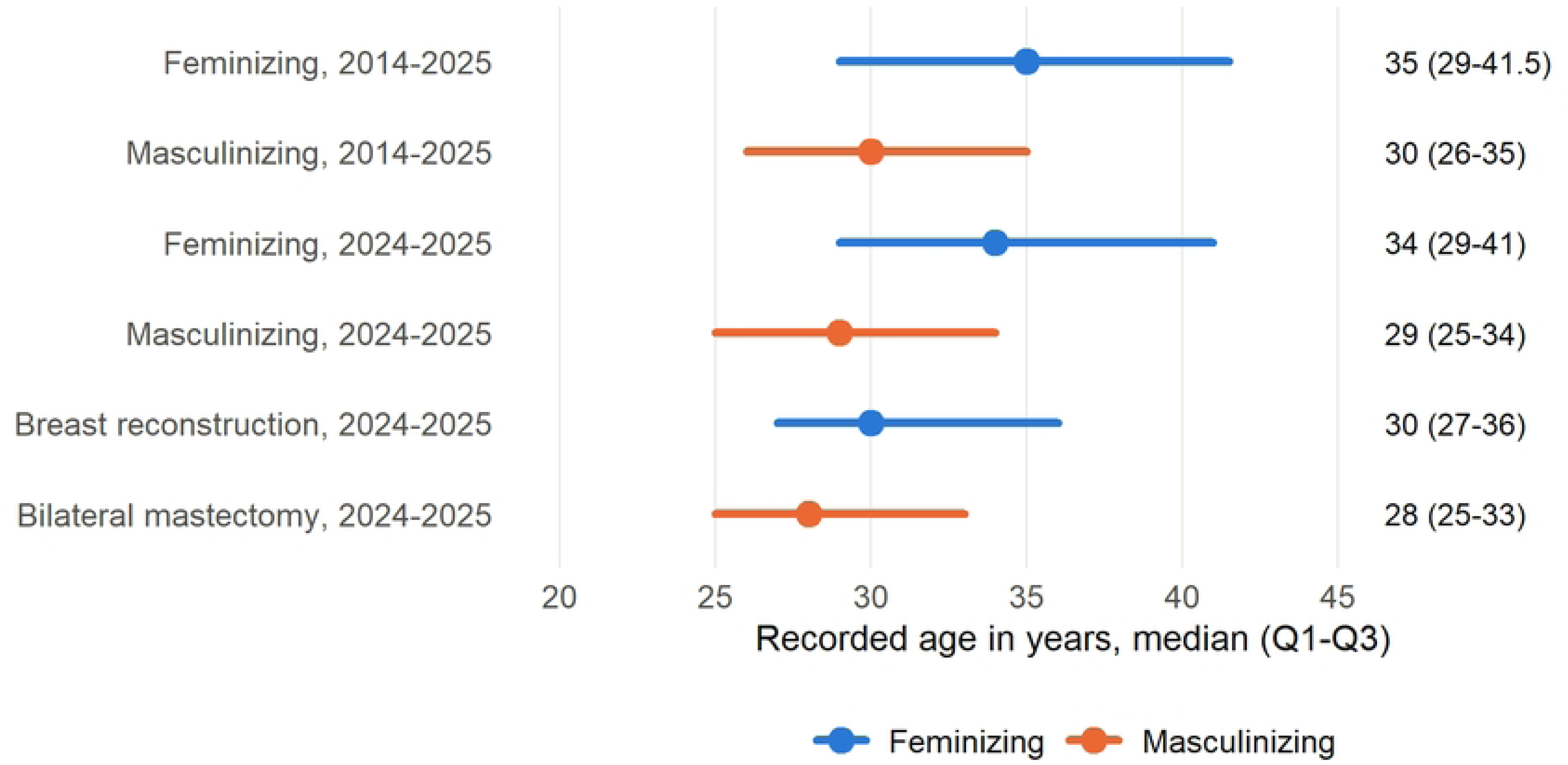
Recorded age of directly classified authorizations, Brazil, 2014-2025. Points show the median and bars the first to third quartiles (Q1-Q3), for each directed group over the whole period and in 2024-2025 and for the two breast and chest procedures in 2024-2025.

### Breast and chest procedures in 2024-2025

In 2024-2025, bilateral mastectomy accounted for 264 of the 302 masculinizing authorizations (87.4%), and the other masculinizing procedures for 38 (12.6%), whereas bilateral breast reconstruction with silicone implants accounted for 118 of the 302 feminizing authorizations (39.1%), and the other feminizing procedures for 184 (60.9%) (Table 3). The median recorded age was 28 years (25-33) for mastectomy and 30 years (27-36) for breast reconstruction, and the brown category was recorded in 198 of 264 (75.0%) and in 75 of 118 (63.6%) of these authorizations. Annual counts of bilateral mastectomy were 24 in 2023, 102 in 2024 and 162 in 2025, and those of breast reconstruction were 12, 34 and 84 in the same years.

**Table 3.** Breast and chest procedures among directly classified authorizations, Brazil, 2024-2025.

| **Procedure (SIGTAP code)** | **Authorizations, n** | **Share of the directed group, n/N (%)** | **Recorded age, median (Q1-Q3)** | **Brown (parda), n (%)** |
|---|---|---|---|---|
| Bilateral mastectomy (0410010197) | 264 | 264/302 (87.4) | 28 (25-33) | 198 (75.0) |
| Bilateral breast reconstruction with silicone implants (0410010200) | 118 | 118/302 (39.1) | 30 (27-36) | 75 (63.6) |

Note. The unit is the hospital admission authorization (AIH). The denominators are the 302 masculinizing and the 302 feminizing authorizations of 2024-2025. The two operations have different indications and eligibility requirements, and their counts do not represent comparable groups of people. Q1, first quartile. Q3, third quartile.

## Discussion

The present study showed that the convergence of feminizing and masculinizing authorizations in 2024-2025, with 302 authorizations in each group, resulted from procedure portfolios of very different composition, since bilateral mastectomy accounted for almost nine in ten masculinizing authorizations, whereas breast reconstruction accounted for about four in ten feminizing authorizations and the remainder was distributed among genital procedures, thyroplasty and other operations. Although the masculinizing genital procedure and neophalloplasty entered the procedure table in 2019 and 2024, all procedures other than mastectomy, including hysterectomy, contributed only 38 of the 302 masculinizing authorizations of the recent period. This observation suggests that the expansion of the coded portfolio had a small direct weight in the convergence and that the growth was concentrated in a single operation. This small contribution may reflect the staged character of masculinizing genital reconstruction and the fact that the national ordinance classified vaginectomy and neophalloplasty as experimental procedures restricted to accredited teaching hospitals [1], which may have delayed their provision. In the feminizing group, the genital procedure remained stable across the three periods, with 151, 138 and 150 authorizations, so that the recent growth of both groups was concentrated in breast and chest surgery. In this sense, a ratio between feminizing and masculinizing counts may be misleading as an indicator of balance in service provision, because hospital authorizations are administrative outputs of coded procedures and do not measure the number of people served, the availability of each procedure, need, waiting time or equity of access.

The annual counts of bilateral mastectomy observed in the present study, 24 in 2023 and 162 in 2025, coincide with those reported in an independent analysis of the same information system [13], which reinforces the reproducibility of the selection. On the other hand, another conference analysis reported 1,268 hospitalizations from January 2014 to November 2025 [14], against the 1,375 authorizations selected here. Since both series start with 24 hospitalizations in 2014, it is possible that the difference arises from the shorter observation window of that analysis, which did not include December 2025, and from differences in procedure groupings, aspects that the available information does not allow to reconcile fully. The predominance of breast and chest surgery among masculinizing authorizations is also consistent with national estimates from the United States, in which breast and chest procedures were the most frequent group of gender-affirming surgery [6,7], although differences in health systems, coverage rules and units of analysis limit direct comparison.

Nevertheless, the concentration of recent growth in one operation suggests that changes in a single line of care, such as the accreditation of new services or the organization of surgical queues, may be sufficient to modify national aggregates, hypotheses that cannot be tested with the present data. In this direction, the number of establishments habilitated for these surgeries doubled in 2023, from five to ten, and from September 2023 the SIGTAP also accepted the habilitation of the national program for reducing elective surgery queues, changes that coincide with the increase in authorizations, although the authorizations could not be attributed to each establishment or program. The decrease in selected authorizations in 2020 and 2021 coincides with the COVID-19 pandemic, when a reduction of 46% in elective procedures performed by the SUS over nine months of 2020 was attributed to the pandemic [26], and with the temporary decline and later resurgence of gender-affirming surgery observed in the United States [27].

The age difference of about five years between the groups was observed both over the whole period and within 2024-2025, which reduces the possibility that it reflects only the pooling of different calendar periods. The difference may be related to the procedure mix, since mastectomy, which predominated among masculinizing authorizations, had a lower median age than breast reconstruction, as well as to eligibility and referral pathways, the organization of establishments and repeat authorizations. It is worth noting that the national ordinance set the minimum age of 21 years and two years of prior follow-up for surgical procedures [1], which may contribute to the concentration of authorizations in young adulthood. In this context, the few records with recorded ages below 21 years, all processed before 2024 and including values incompatible with any surgical indication, may be more plausibly explained by errors in the age field than by surgery outside the eligibility rule, a possibility that could only be confirmed by audit of the original records.

The predominance of mastectomy among masculinizing authorizations and its lower median age may be related to the place of the chest in the experience of trans men. In a large community survey, chest binding with commercial binders, elastic bandages or tape was associated with at least one negative physical outcome in more than 97% of participants [28], and, in a Brazilian study, masculinizing mammoplasty was associated with greater satisfaction with chest appearance and with less frequent binding [29,30], findings that may help explain an earlier search for this operation. In the feminizing group, the breast development produced by hormone therapy is modest in many cases [31], which may contribute to the continued demand for breast reconstruction, while the stability of genital procedures across the three periods is compatible with evidence that the desire for each gender-affirming intervention is heterogeneous and that part of trans people do not wish to undergo surgery [32,33]. These interpretations refer to needs and preferences that billing records do not measure and would require patient-reported data to be confirmed.

In relation to race/color, the larger share of the brown category among masculinizing authorizations, 61.0% against 42.6%, was observed in a comparison pooled over the whole period and not adjusted for period, procedure or establishment. Considering that the masculinizing group was concentrated in 2024-2025 and in mastectomy, this difference may reflect changes in the regional distribution of accredited services and in the composition of the recent series, since an independent analysis reported the largest number of mastectomies in the Northeast region [13]. This possibility is supported by the observation that, within 2024-2025, the brown category was recorded in 75.0% of mastectomies and in 63.6% of breast reconstructions, both above the pooled share of the feminizing group, although these procedure-specific figures were not adjusted for establishment. In addition, in the 2019 National Health Survey, private health insurance covered 38.8% of white people, against 21.4% of black and 20.1% of brown people [34], so that a greater dependence on the SUS among black and brown people may contribute to their larger share among publicly financed authorizations, a relationship that was not examined among trans people. For this reason, the finding was not interpreted as a racial disparity at the person level. It should also be noted that race/color recorded in hospital authorizations depends on recording practices that were not characterized and that the small number of authorizations recorded as black, 64 and 17, limits the analysis of this population, although black and brown people together accounted for the majority of authorizations in both groups. The interpretation of these categories requires attention to the local social context, as studies with patient-reported data have shown [17].

From the perspective of global public health, the findings suggest that administrative surveillance of publicly funded gender-affirming surgery may document the volume and composition of billed care and raise questions for service planning, provided that indicators are disaggregated by procedure and that code definitions are versioned over time. The near-universal recording of a principal diagnosis in category F64 (97.7%) is consistent with the SIGTAP rule that accepts only F64.0, transsexualism, as the principal diagnosis for these procedures [23], a category that the ICD-10, still used in the SIH/SUS, places among mental disorders. The ICD-11 reconceptualized this category as gender incongruence and moved it to a chapter on sexual health [35,36], so that the adoption of the ICD-11 by the information systems may require a revision of this rule that preserves access to care. The investigation of access, continuity of care and equity would require person-linked records, waiting-list denominators and patient-reported experiences [16]. Relative to previous Brazilian descriptions of policy and surgical volumes [2–5,13,14], the contribution of the present study lies in the joint description of historically eligible procedure codes, recorded age and race/color and the procedure composition behind the recent convergence of counts, together with the reproduction of all aggregate results from the original public files.

The present study was limited to administrative episodes, and an authorization may recur for the same person, since no validated person-level linkage was available, while the direction of the procedure does not measure self-reported gender identity, sex assigned at birth, clinical indication or preference. Fields such as date of birth, sex and municipality of residence could support a probabilistic approximation of unique persons, an approach that was not adopted because it has not been validated for these records and would increase the risk of re-identification in small groups. For the same reason, it was not possible to estimate how many people completed the successive stages of masculinizing genital surgery. Eligibility rules, financing and recording practices changed over 2014-2025, the periods compared have unequal durations, and shifts in the feminizing share cannot be read as population rates or as effects of policy. The same-period age comparison reduces historical heterogeneity but does not adjust for procedure or establishment, and race/color findings are pooled over the whole period, not adjusted and not confirmed as self-declared. The independent replay reproduced all aggregate results from the original files but cannot verify the clinical validity of the coding or the total care delivered, and the agreement with the national establishment register refers only to administrative geography, without information on place of residence or travel.

However, it should be noted that the use of all public files of the period, the historical versioning of procedure codes and the independent reproduction of the results allow other researchers to repeat and extend the analysis with the same public sources.

## Conclusions

The study made it possible to identify that, in the SUS, the equal numbers of directly feminizing and masculinizing surgical authorizations in 2024-2025 corresponded to different procedure portfolios, with bilateral mastectomy accounting for 87.4% of masculinizing authorizations and breast reconstruction for 39.1% of feminizing authorizations. It was also observed that a difference of about five years in median recorded age persisted within the recent period, whereas the race/color differences remained pooled and unadjusted. These findings support procedure-specific and historically versioned monitoring of publicly funded gender-affirming surgery and indicate that conclusions about gender identity, individual demand or equity of access would require data on people and on their experiences of care.

## Data Availability

All data underlying the findings are publicly available. The reduced files of the Hospital Information System of the Brazilian Unified Health System (SIH/SUS) are disseminated by DATASUS at ftp://ftp.datasus.gov.br/dissemin/publicos/SIHSUS/200801_/Dados/ (files RD[UF][YYMM].dbc, January 2014 to December 2025), the monthly procedure tables (SIGTAP) at ftp://ftp2.datasus.gov.br/pub/sistemas/tup/downloads/ and the habilitation files of the National Registry of Health Establishments (CNES) at ftp://ftp.datasus.gov.br/dissemin/publicos/CNES/200508_/Dados/HB/. The analysis code, the historical procedure crosswalk, the notebook that counts the habilitated establishments, the aggregate results reported in the article, the software environment and a checksum manifest are deposited in Zenodo (https://doi.org/10.5281/zenodo.24117844) under the Creative Commons Attribution 4.0 International (CC BY 4.0) license. The event-level extract derived by the authors from the public files is not redistributed, because combinations of rare procedures and social attributes may raise a risk of indirect identification, and it can be regenerated in full from the public DATASUS files with the deposited code.

ftp://ftp.datasus.gov.br/dissemin/publicos/CNES/200508_/Dados/HB/

ftp://ftp2.datasus.gov.br/pub/sistemas/tup/downloads/

ftp://ftp.datasus.gov.br/dissemin/publicos/SIHSUS/200801_/Dados/

https://doi.org/10.5281/zenodo.24117844

## Acknowledgments

None.

## Supporting information

S1 Appendix. Data provenance, historical procedure crosswalk, selection flow, quality checks and disclosure controls.

S2 Protocol. Analysis protocol documented retrospectively, with definitions and deviations. S3 Checklist. STROBE and RECORD checklist with the location of each item in the manuscript.

